# In schizophrenia, chronic fatigue syndrome- and fibromyalgia-like symptoms are driven by breakdown of the paracellular pathway with increased zonulin and immune activation-associated neurotoxicity

**DOI:** 10.1101/2021.05.09.21256897

**Authors:** Michael Maes, Laura Andrés-Rodríguez, Aristo Vojdani, Sunee Sirivichayakul, Decio S. Barbosa, Buranee Kanchanatawan

## Abstract

**Background:** A meaningful part of schizophrenia patients suffer from physiosomatic symptoms (formerly named psychosomatic) which are reminiscent of chronic fatigue syndrome and fibromyalgia (FF) and are associated with signs of immune activation and increased levels of tryptophan catabolites (TRYCATs).

**Aims:** To examine whether FF symptoms in schizophrenia are associated with breakdown of the paracellular pathway, zonulin, lowered natural IgM responses to oxidative specific epitopes (OSEs); and whether FF symptoms belong to the behavioral-cognitive-physical-psychosocial-(BCPS)-worsening index consisting of indices of a general cognitive decline (G-CoDe), symptomatome of schizophrenia, and quality of life (QoL)-phenomenome.

**Methods:** FF symptoms were assessed using the Fibromyalgia and Chronic Fatigue Rating scale in 80 schizophrenia patients and 40 healthy controls and serum cytokines/chemokines, IgA levels to TRYCATs, IgM to OSEs, zonulin and transcellular/paracellular (TRANS/PARA) molecules were assayed using ELISA methods.

**Results:** A large part (42.3%) of the variance in the total FF score was explained by the regression on the PARA/TRANS ratio, pro-inflammatory cytokines, IgM to zonulin, IgA to TRYCATs (all positively) and IgM to OSEs (inversely). There were highly significant correlations between the total FF score and G-CoDe, symtopmatome, QoL phenomenome and BCPS-worsening score. FF symptoms belong to a common core shared by G-CoDe, symtopmatome, and QoL phenomenome. Discussion: The physio-somatic symptoms of schizophrenia are driven by various pathways including increased zonulin, breakdown of the paracellular tight-junctions pathway, immune activation with induction of the TRYCAT pathway, and consequent neurotoxicity. It is concluded that FF symptoms are part of the phenome of schizophrenia and BCPS-worsening as well.

## Introduction

Recently, we developed a new method, namely nomothetic network psychiatry, to model major psychiatric disorders including schizophrenia and major depressive and bipolar disorder based on causome, AOP (adverse outcome pathways), and phenome data (Maes et al., 2020b; 2021a; Stoyanov and Maes, 2021). Applied to schizophrenia, we constructed a new reliable and cross-validated, data-driven model, which assembles the effects of risk (causome) and resilience (protectome) features, different AOPs (neuro-immune pathways), cognitome (i.e., the aggregate of neuro-cognitive impairments), symptomatome (i.e., the aggregate of all symptoms of schizophrenia), and phenomenome (i.e., the self-experience of illness features by the subjects) features (Maes et al., 2020b).

We conceptualized the symptomatome of schizophrenia as the first factor extracted from newly developed scores of psychosis, hostility, excitation, mannerism (PHEM), negative symptoms, psychomotor retardation (PMR), and formal thought disorders (FTD) (Almulla et al., 2020b; Al-Hakeim et al., 2020). We conceptualized the cognitome as the first factor extracted from two neuropsychological batteries, namely the Cambridge Neuropsychological Test Automated Battery and the Consortium to Establish a Registry for Alzheimer’s disease and including test scores reflecting disorders in executive functions, strategy use, attention set-shifting, visual sustained attention, rule acquisition, emotional recognition, semantic and episodic memory, and recall (Maes and Kanchanatawan et al., 2021). These factor scores reflect the severity of the General Cognitive Decline (G-CoDe) in schizophrenia (Maes and Kanchanatawan et al., 2021). The phenomenome of schizophrenia was conceptualized as the first factor extracted from self-rated scores of physical health, psychological health, and social and environmental responsible behaviors as scored with the World Health Organization Quality of Life Instrument-Abbreviated version (WHO-QoL-BREF) (Kanchanatawan et al., 2019). Moreover, we also constructed a new index which captures the severity of deterioration or the deficit in schizophrenia namely the behavioral-cognitive-physical-psycho-social-(BCPS)-worsening index conceptualized as the first factor extracted from the symptomatome, cognitome (G-CoDe), and QoL-phenomenome (Maes et al., 2021b).

Recently, we reported that a meaningful part of schizophrenia patients suffer from physiosomatic symptoms (formerly named psychosomatic symptoms) which are reminiscent of chronic fatigue syndrome and fibromyalgia (Kanchanatawan et al 2017; Almulla et al., 2020a; Mousa et al., 2021). We assessed these symptoms using the Fibromyalgia and Chronic Fatigue Rating (FF) Scale, including fatigue, muscle tension, cramps and pain, a flu-like malaise, headache, and autonomic and cognitive symptoms (Kanchanatawan et al 2017; Almulla et al., 2020a; Mousa et al., 2021). Moreover, the FF symptoms were strongly correlated with PHEM and negative symptoms, PMR, and FTD (Almulla et al., 2020a). The total FF score loaded highly on the same latent vector, which was extracted from these typical schizophrenia symptoms, suggesting that FF symptoms are another manifestation of the symptomatome of that illness (Almulla et al., 2020a). Furthermore, FF symptoms were strongly associated with impairments in semantic and episodic memory, working memory, executive functions, spatial planning, paired associative learning, and attention (Kanchanatwan et al., 2017; Almulla et al., 2020a). In another study, we found that health-related (HR)-Qol in schizophrenia was best predicted by severity and anxiety and FF symptoms and that these symptoms were more important than psychosis and negative symptoms in predicting a lowered HR-QoL (Kanchanatawan et al., 2019). Nevertheless, there are no data whether FF symptoms also contribute to the BCPS-worsening index.

There is now also evidence that the symptomatome, cognitome, Qol-phenomenome, and the BCPS-index are strongly and positively associated with different AOPs, namely with a) biomarkers of immune activation including increased levels of pro-inflammatory cytokines/chemokines such as interleukin-6 (IL-6), MIF-1, tumor necrosis factor (TNF)-α, IL-1β, and CCL11 (or eotaxin); b) activation of the tryptophan catabolite (TRYCAT) pathway as assessed with IgA responses to diverse TRYCATs; c) breakdown of the paracellular gut pathway with increased levels of zonulin; d) increased neurotoxicity as measured with the above cytotoxic cytokines/chemokines coupled with biomarkers of oxidative stress including lipid hydroperoxides (LOOH), malondialdehyde (MDA), and advanced oxidation protein products (AOPP); and e) increased levels of lipopolysaccharides of Gram-negative bacteria. Moreover, we found also negative association between the above clinical features of schizophrenia and natural IgM to oxidatively specific epitopes (OSEs), including MDA and azelaic acid, which generally have immune-regulatory properties (Maes et al., 2021b).

Importantly, increased expression of FF symptoms in schizophrenia is associated signs of immune activation, including elevated IgA responses to neurotoxic TRYCATs such as picolinic acid (PA), xanthurenic acid (XA), and 3-OH-kynurenine (3HK) (Kanchanatawan et al., 2019), and increased levels of IL-6, CCL11, and IL-1β (Mousa et al., 2021; Almulla et al., 2020a). Nevertheless, there are no data whether FF symptoms in schizophrenia are associated with indicants of the paracellular gut pathway, increased levels of zonulin and neurotoxicity, and lowered levels of protective IgM to OSEs.

Hence, the present study was conducted to a) examine whether FF symptoms in schizophrenia are associated with breakdown of the paracellular pathway as assessed with IgM responses to paracellular / transcellular molecules, IgM responses to zonulin and OSEs, immune activation and neurotoxicity indices, and IgA responses to TRYCATs; and b) examine whether FF symptoms belong to the BCPS-worsening index and, consequently, constitute another manifestation of the BCPS-worsening in schizophrenia.

## Subjects and Methods

### Participants

This is a case-control study which recruited Thai participants, both men and women aged between 18 and 65 years old. We recruited schizophrenia outpatients at the Department of Psychiatry, Faculty of Medicine, Chulalongkorn University, Bangkok, Thailand and normal controls from the same catchment area (Bangkok, Thailand). The patients complied with the diagnostic criteria for schizophrenia according to DSM-IV-TR criteria. They were all in a clinically stable phase of illness for at least one year and were treated with the same antipsychotic maintenance treatment during that period. The healthy controls were healthy individuals, and they were recruited by word of mouth as friends or family of staff members and friends of the patients.

Schizophrenia patients were excluded when they showed a lifetime or current diagnosis of axis-I DSM-IV-TR or DSM-5 psychiatric disorders, including psycho-organic disorders, bipolar disorder, major depressive disorder, and substance use disorders (all except nicotine dependence). We excluded controls with any lifetime/current DSM-IV-TR / DSM-5 axis 1 diagnoses of psychiatric illness and/or with a positive family history of psychosis. Controls and patients were excluded when they showed: a) systemic (auto)immune or inflammatory illness, such as rheumatoid arthritis, psoriasis, diabetes, chronic obstructive pulmonary disease, and inflammatory bowel disease; b) neuroinflammatory or neurodegenerative disorders including Alzheimer’s and Parkinson’s disorder, stroke and multiple sclerosis; c) lifetime (or current) treatment with immunomodulatory drugs such as methotrexate and glucocorticoids; d) recent use (last 6 months) of therapeutical dosages of antioxidants or omega-3 supplements; and e) alcohol abuse. We also excluded pregnant or lactating women.

All controls and patients as well as their guardians (parents or other close family members of the patients) gave written informed consent prior to participation in this study. The study was conducted according to Thai and international ethics and privacy laws. Approval for the study was obtained from the Institutional Review Board of the Faculty of Medicine, Chulalongkorn University, Bangkok, Thailand (No 298/57), which is in compliance with the International Guideline for Human Research protection as required by the Declaration of Helsinki, The Belmont Report, CIOMS Guideline and International Conference on Harmonization in Good Clinical Practice (ICH-GCP).

### Methods

A senior psychiatrist (BK) specialized in schizophrenia research completed a semi-structure interview with socio-demographic and clinical data. We employed the Mini-International Neuropsychiatric Interview (M.I.N.I.) in a validated Thai translation (Kittirathanapaiboon and Khamwongpin, 2005) to make the diagnosis of schizophrenia and to exclude individuals with other psychiatric disease. The same senior psychiatrist rated the Fibromyalgia and Chronic Fatigue Syndrome Rating (FF) scale (Zachrisson et al. 2002) in all participants. The sum of all 12 symptoms was employed as an index of severity of the FF symptomatome. The 12 items of the FF scales are “FF1: muscle pain, FF2: muscular tension, FF3: fatigue, FF4: concentration difficulties, FF5: failing memory, FF6: irritability, FF7: sadness, FF8: sleep disturbances, FF9: autonomic disturbances, FF10: irritable bowel, FF11: headache, and FF12: a flu-like malaise” (Kanchanatawan et al., 2017). We computed the sum of selected physiosomatic FF symptoms, namely FF1 + FF2 + FF3 + FF9 + FF10 + FF11 + FF12 thereby deleting the cognitive and affective FF symptoms from the FF physiosomatic score.

To compute the severity of the symptomatome of schizophrenia, we assessed the Brief Psychiatric Rating Scale (BPRS) (Overall and Gorham, 1962), the Positive and Negative Syndrome Scale (PANSS) (Kay et al., 1986), the Scale for the Assessment of Negative Symptoms (SANS) (Andreasen et al., 1989), and the Hamilton Depression (HDRS) (Hamilton, 1960). Using the items of the BPRS, PANSS, SANS, and HDRS we computed z unit-weighted based composite scores reflecting the severity of psychosis, hostility, excitation, mannerism, psychomotor retardation (PMR) and formal thought disorders (Almulla et al., 2020a; Al-Hakeim et al., 2020; Maes and Kanchanatawan, 2021). Consequently, we extracted the first PC from these symptom domain scores and, therefore, this score reflects the severity of the symptomatome of schizophrenia. The same day, all participants completed the WHO-QoL-BREF (WHO, 1993), a rating scale which assesses four HR-QoL domains, namely physical and psychological health, relationships, and environment. To compute the QoL-phenomenome we extracted the first PC from these 4 domain scores as explained previously (Maes and Kanchanatawan, 2021).

The same day, a well-trained clinical research assistant assessed four CERAD tests, i.e., the Verbal Fluency Test (VFT), Word List Memory (WLM), WLM True Recall, and the Mini Mental State Examination (MMSE), and CANTAB Research Suite Tests, i.e., spatial working memory between errors (SWM-BE) and SWM strategy (SWM-STR), rapid visual information process test A’ Prime (RVP_A), and paired-association learning total errors adjusted (PAL-TEA), One touch stockings of Cambridge probability solved on first choice (OTS-PSOFC), Emotional recognition test mean overall response latency (ERT-MORL) and the Intra/extradimensional set shifting total errors adjusted (IED_TEA) (CERAD, 1986; CANTAB, 2018). As explained previously, we extracted the first LV from these different test scores and this first PC reflects the G-CoDe (Maes and Kanchanatawan, 2021). Finally, we have extracted the first PC from the symptomatome, G-CoDe, and the QoL-phenomenome factor scores which, therefore, reflects the BCPC-worsening index (Maes et al., 2021b). Consequently, we have binned the BCPS-worsening index of the patients into two groups using the median-split method thereby forming two patient groups, namely patients with a low (SCZ-BCPS) and a more severe (SCZ+BCPS) worsening index.

DSM-IV-TR criteria were used to make the diagnosis of tobacco use disorder (TUD). Body mass index (BMI) was computed as body weight (in kg) divided by body length (in meter) squared.

### Assays

Fasting blood was sampled at 8.00 a.m. and serum and plasma Eppendorfs were frozen at −80 °C until thawed for assay. Blood samples of controls and patients were taken at the same time, using exactly the same procedures. Each set of assays was performed in the same run by the same operator who was blinded to the clinical results. Serum levels of IL-6, IL-10, TNF-α, MIP-1, CCL11, and the soluble IL-1 receptor antagonist (sIL-1RA) (R&D Systems, Inc, Minneapolis, MN, USA) were assayed employing the Bio-Plex^®^ 200 System (Bio-Rad Laboratories, Inc.) [22,24]. AOPP was assayed in a microplate reader (EnSpire, Perkin Elmer, USA) at a wavelength of 340 nm. LOOH was assayed by chemiluminescence in a Glomax Luminometer (TD 20/20), in the dark, at 30 °C for 60 min. MDA was assayed through complexation with two molecules of thiobarbituric acid and high-performance liquid chromatography (HPLC Alliance e2695, Waters**’**, Barueri, SP, Brazil). Indirect ELISA tests were used to measure IgM directed against conjugated azelaic acid, oleic acid, MDA, and phosphatidylinositol (Pi) (Maes et al., 2021b). Consequently, we extracted the first factor from those 4 optical density values, which reflects the IgM protectome. As described previously, ELISA tests were employed to assay the plasma titers of IgA directed against XA (Acros), PA (Acros), 3HK (Sigma), anthranilic acid (AA) (Akros), and kynurenic acid (KA) (Akros) linked to 20 mg BSA (ID Bio) (Kanchanatawan et al., 2019). We measured the optical densities (ODs) at 450 nm using Varioskan Flash (Thermo Scientific). In order to compute the paracellular/transcellular (PARA/TRANS) leaky gut ratio we computed a z unit-weighted composite score as z (z zonulin + z occludin + z E-cadherin) – z (z talin - z actin - z vinculin). Zonulin, occludin, E-cadherin, talin and vinculin were purchased from Bio-Synthesis® (Lewisville, TX, USA) and Abcam® (Cambridge, MA, USA) and actin from Sigma-Aldrich® (St. Louis, MO, USA). Diluted IgM was added to each well of the microtiter plate and optical densities were determined as described in Maes et al. (2019a). The assays of IgA responses directed against *Hafnia alvei, Klebsiella pneumonia, Morganella morganii, Pseudomonas aeruginosa*, and *Pseudomonas putida. w*ere conducted using ELISA techniques as described by Roomruanwong et al. (2018). Consequently, we computed a z unit-weighted composite score summing up the 5 z transformed values of IgA to the 5 Gram-negative bacteria (IgA LPS). This score reflects plasma LPS load following increased gut permeability or leaky gut.

Based on these assays we computed a number of z composite scores: a) z score of PA (zPA)+ z3HK + zXA – zAA – zKA, reflecting the noxious / generally more protective TRYCAT ratio (Kanchanatawan et al., 2019); b) sum of 4 cytokines which are increased during immune responses (4 cytokines index) as sum a z scores of zIL-6 + zTNF-α + zIL10 + zMIP-1; c) the neurotoxicity index was computed as zTNF-α + zIL-6 + zMIP + z(zXA+PA+3HK) + zLOOH + zMDA + zAOPP. CCL11 was not included in this index to allow for the additional effects of this chemokine; and d) the MITOTOX index was computed as zTNF-α + zIL-6 + zCCL11 + zTRYCAT ratio + zLOOH + zMDA + zAOPP + zIgA LPS (Maes et al., 2020a; 2021b).

### Statistics

We employed analysis of variance (ANOVA) to check differences in scale variables between groups and analysis of contingency tables (X^2^-test) or Fisher’s exact probability test to check associations between categorical variables. Associations between two scale variables were assessed using Pearson’s product moment correlation coefficients. In order to correct for type 1 error due to multiple comparisons we employed a p correction for false discovery rate (FDR). Multiple regression analysis was employed to assess the significant predictors of the FF scores, and we employed the R^2^ value as an estimate of the effect size. We allowed for the effects of sex, age, education, and BMI when examining the effects of biomarkers on FF scores and significant background variables were included in the final model. All regression analyses were checked for multicollinearity. We used principal component analysis to extract PCs from a data set and latent vectors (LV) were considered to be reliable when the explained variance was > 50.0% and all variables loaded highly (>0.66) on the first PC. The adequacy for factorization was checked with the Kaiser-Meyer-Olkin (KMO) test and Bartlett’s test of sphericity. Statistical analyses were performed using IBM SPSS Windows version 25. Tests were 2-tailed, and an alpha level of 0.05 indicated a statistically significant effect.

## Results

### Sociodemographic and clinical data

**Table 1** shows the sociodemographic data in schizophrenia patients divided into those with and without an increased BCPS-worsening index and normal controls. There were no significant differences in age, BMI, nicotine dependence, between the three study groups. There were significantly more males in the schizophrenia group than in controls. The unemployment rate increased from controls → SCZ-BCPS → SCZ+BCPS. The total FF and FF physiosomatic, symptomatome, and BCPS scores were significantly different between the three study groups and increased from controls → SCZ-BCPS → SCZ+BCPS. The G-CoDe and QoL phenomenome decreased from controls to SCZ-BCPS to SCZ+BCPS. These differences in clinical indices remained significant after FDR p correction.

**Table 1.** Demographic and clinical data in normal controls and schizophrenia (SCZ) patients divided into those with and without a severe BCPS (behavioral-cognitive-physical-psychosocial)-worsening index.

| Variables | Controls | SCZ - BCPS | SCZ + BCPS | F/X<sup>2</sup> | df | p |
| --- | --- | --- | --- | --- | --- | --- |
| Age (years) | 37.4 (12.8) | 40.3 (11.2) | 41.9 (10.9) | 1.55 | 2/117 | 0.217 |
| Sex (M/F) | 30/10 | 19/20 | 18/23 | 9.13 | 3 | 0.010 |
| BMI (kg/m<sup>2</sup>) | 24.0 (4.3) | 25.5 (5.7) | 23.5 (4.4) | 1.75 | 2/112 | 0.179 |
| Education (years) | 14.3 (4.9)<sup>C</sup> | 13.4 (3.8)<sup>C</sup> | 11.3 (4.3)<sup>A,B</sup> | 5.12 | 2/117 | 0.007 |
| Employment (N/Y) | 4/36<sup>B,C</sup> | 15/24<sup>A,C</sup> | 31/10<sup>A,B</sup> | 36.10 | 2 | <0.001 |
| Nicotine dependence (N/Y) | 38/2 | 36/3 | 39/2 | FEPT | - | NS |
| FF total score | 1.40 (3.35)<sup>B,C</sup> | 9.03 (6.31)<sup>A,C</sup> | 15.78 (8.25)<sup>A,B</sup> | 52.43 | 2/116 | <0.001 |
| FF physiosomatic score | 0.73 (1.93)<sup>B,C</sup> | 4.92 (4.18)<sup>A,C</sup> | 8.88 (5.26)<sup>A,B</sup> | 41.11 | 2/116 | <0.001 |
| Symptomatome (z score) | -0.902 (0.032)<sup>B,C</sup> | -0.205 (0.551)<sup>A,C</sup> | 1.076 (0.803)<sup>A,B</sup> | 127.78 | 2/117 | <0.001 |
| General Cognitive decline (z score) | 0.797 (0.741)<sup>B,C</sup> | 0.072 (0.657)<sup>A,C</sup> | -0.846 (0.810)<sup>A,B</sup> | 50.12 | 2/117 | <0.001 |
| QoL-phenomenome (z score) | -0.793 (0.598)<sup>B,C</sup> | 0.134 (0.847)<sup>A,C</sup> | -0.901 (0.683)<sup>A,B</sup> | 57.78 | 2/117 | <0.001 |
| BCPS-worsening score (z score) | -0.956 (0.335)<sup>B,C</sup> | -0.256 (0.495)<sup>A,C</sup> | 1.176 (0.486)<sup>A,B</sup> | 242.52 | 2/117 | <0.001 |
| CCL11 mg/dL | 129.6 (54.1)<sup>B,C</sup> | 196.7 (90.2)<sup>A</sup> | 225.0 (108.9)<sup>A</sup> | 12.62 | 2/117 | <0.001 |
| 4 cytokines (z score) | -0.564 (0.678)<sup>B,C</sup> | 0.027 (0.861)<sup>A,C</sup> | 0.524 (1.105)<sup>A,B</sup> | 14.81 | 2/117 | <0.001 |
| IgA TRYCATs (z score) | -0.695 (0.655)<sup>B,C</sup> | 0.225 (0.888)<sup>A</sup> | 0.464 (1.027)<sup>A</sup> | 19.86 | 2/117 | <0.001 |
| Immune activation index (z score) | -0.746 0.755 B,C | 0.088 0.718 A,C | 0.643 0.970 A,B | 19.76 | 2/117 | <0.001 |
| Neurotoxicity index (z score) | -0.509 0.834 B,C | -0.068 0.823 A,C | 0.561 1.034 A,B | 14.35 | 2/117 | <0.001 |
| MITOTOX index (z score) | -0.768 0.684 B,C | 0.045 0.786 A,C | 0.649 0.956 A,B | 28.30 | 2/117 | <0.001 |
| IgM protectome (z score) | 0.263 (0.833) C | 0.004 (0.937) | -0.261 1.151 A | 2.87 | 2/117 | 0.061 |
| IgM PARA/TRANS (z score) | -0.247 (0.900) C | -0.183 (0.918) C | 0.412 1.057 A,B | 5.63 | 1/114 | 0.005 |
| IgM zonulin (z score) | -0.123 0.849 | 0.148 1.138 | -0.028 0.994 | 0.73 | 2/114 | 0.486 |

All results are shown as mean (SD); <sup>A,B,C</sup>: pairwise comparisons among group means.
FEPT: Fisher's exact probability test; BMI: body mass index.
FF: The Fibromyalgia and Chronic Fatigue Rating (FF) scale scores.
PARA/TRANS: paracellular / transcellular leaky gut ratio; TRYCATs: tryptophan catabolites.

CCL11 and IgA TRYCATs were significantly higher in schizophrenia than in controls. The 4 cytokines, immune activation, neurotoxicity, and MITOTOX indices were significantly different between the three subgroups and increased from controls → SCZ-BCPS → SCZ+BCPS. The IgM protectome was significantly lower in SCZ+BCPS than in controls, and IgM PARA/TRANS significantly higher in SCZ+BCPS than in the two other groups. The differences in biomarkers remained significant after p correction. The differences in FF scores, clinical indices, and biomarkers were not significantly affected by the drug state of the patients (even without FDR p correction), namely antidepressants n=28, risperidone n=34, olanzapine n= 5, quetiapine n=4, clozapine n=10, haloperidol n=12, perphenazine n=21, chlorpromazine n=5. There were no differences in the drug state (or chlorpromazine equivalents) between SCZ-BCPS and SCZ+BCPS patients.

### Intercorrelation matrix

**Table 2** shows the correlations (after FDR p correction) between both FF scores and clinical and biomarker data. Both the total and physiosomatic FF scores were significantly associated with the symptomatome, BCPS-worsening score, CCL11, IgA TRYCATs, and 4 cytokines, immune activation, neurotoxicity, and MITOTOX indices, the IgM TRANS/PARA ratio and zonulin.

**Table 2.** Correlation coefficients between the total and physiosomatic scores of the Fibromyalgia and Chronic Fatigue Rating (FF) scale or the first factor extracted from the total FF score and the BCPS (behavioral-cognitive-physical-psychosocial)-worsening index (BCPS+FF) and clinical indices and biomarkers of schizophrenia.

| Domains | In controls and SCZ combined |  |  |
| --- | --- | --- | --- |
|  | FF total score | FF physiosomatic score | BCPS+FF |
| Symptomatome | 0.635 | 0.608 | - |
| General-Cognitive Decline (G-CoDe) | -0.527 | -0.526 | - |
| Quality of Life (QoL)-phenomenome | -0.638 | -0.627 | - |
| Behavioral-Cognitive-Physical-Psychosocial-(BCPS)-worsening score | 0.732 | 0.720 | - |
| CCL11 (eotaxin) | 0.373 | 0.365 | 0.394 |
| 4 cytokines | 0.406 | 0.370 | 0.450 |
| IgA TRYCATs | 0.395 | 0.380 | 0.400 |
| Immune activation index | 0.406 | 0.370 | 0.545 |
| Neurotoxicity index | 0.260 | 0.239 | 0.415 |
| MITOTOX index | 0.455 | 0.425 | 0.569 |
| IgM protectome | -0.173 (NS) | -0.131 (NS) | -0.244 * |
| IgM paracellular/transcellular ratio (leaky gut) | 0.312 | 0.310* | 0.298 |
| IgM zonulin | 0.189* | 0.223* | 0.048 |

All significant at $p < 0.0005$ ; except \*: significant at $p < 0.05$ (all after False Discovery Rate p-correction); NS: not significant.

### Results of multiple regression analysis

**Table 3**, regression #1 shows that 42.3% of the variance in the total FF score could be explained by the regression on IgM PARA/TRANS, CCL11, IgM zonulin, IgA TRYCATs, and 4 cytokines index (all positively) and IgM protectome (inversely). Entering the G-CoDe in the same regression analysis (regression #2) showed that 44.5% of the variance in the total FF score could be explained by G-CoDe, IgM PARA/TRANS, 4 cytokines index, and IgM zonulin (all positively), and IgM protectome (inversely). IgM zonulin showed (in both regressions) the strongest impact. **Figure 1** shows the partial regression of the total FF score on IgM zonulin after considering the effects of all other variables shown in regression #2.

**Table 3.** Results of multiple regression analyses with the Fibromyalgia and Chronic Fatigue (FF) Rating scale scores as dependent variables and biomarkers as explanatory variables while allowing for the effects of age, sex, and education.

| Dependent variables | Explanatory variables | $\beta$ | t | p | F<sub>model</sub> | df | p | R<sup>2</sup> |
| --- | --- | --- | --- | --- | --- | --- | --- | --- |
| #1. FF total score | Model |  |  |  | 12.71 | 6/104 | <0.001 | 0.423 |
|  | IgM PARA/TRANS | 0.188 | 2.43 | 0.017 |  |  |  |  |
|  | CCL11 | 0.249 | 3.13 | 0.002 |  |  |  |  |
|  | IgM zonulin | 0.391 | 3.71 | <0.001 |  |  |  |  |
|  | IgM protectome | -0.263 | -2.30 | 0.024 |  |  |  |  |
|  | IgA TRYCATs | 0.201 | 2.43 | 0.017 |  |  |  |  |
|  | 4 cytokines | 0.177 | 2.06 | 0.042 |  |  |  |  |
| #2. FF total score | Model |  |  |  | 17.50 | 5/105 | <0.001 | 0.445 |
|  | G-CoDe | -0.378 | -4.86 | <0.001 |  |  |  |  |
|  | IgM PARA/TRANS | 0.190 | 2.56 | 0.012 |  |  |  |  |
|  | 4 cytokines | 0.167 | 2.02 | 0.045 |  |  |  |  |
|  | IgM zonulin | 0.409 | 4.04 | <0.001 |  |  |  |  |
|  | IgM protectome | -0.328 | -3.05 | 0.003 |  |  |  |  |
| #3. FF physiosomatic score | Model |  |  |  | 13.16 | 5/105 | <0.001 | 0.385 |
|  | IgM zonulin | 0.415 | 3.85 | <0.001 |  |  |  |  |
|  | IgM protectome | -0.272 | -2.39 | 0.019 |  |  |  |  |
|  | IgA PARA/TRANS | 0.194 | 2.43 | 0.017 |  |  |  |  |
|  | CCL11 | 0.284 | 3.55 | 0.001 |  |  |  |  |
|  | IgA TRYCATs | 0.240 | 2.91 | 0.004 |  |  |  |  |
| #4. FF physiosomatic score | Model |  |  |  | 12.54 | 4/106 | <0.001 | 0.321 |
|  | G-CoDe | -0.354 | -4.17 | <0.001 |  |  |  |  |
|  | IgM zonulin | 0.396 | 3.60 | <0.001 |  |  |  |  |
|  | IgA TRYCATs | 0.181 | 2.06 | 0.042 |  |  |  |  |
|  | IgM protectome | -0.237 | -2.05 | 0.043 |  |  |  |  |

**Figure 1.**
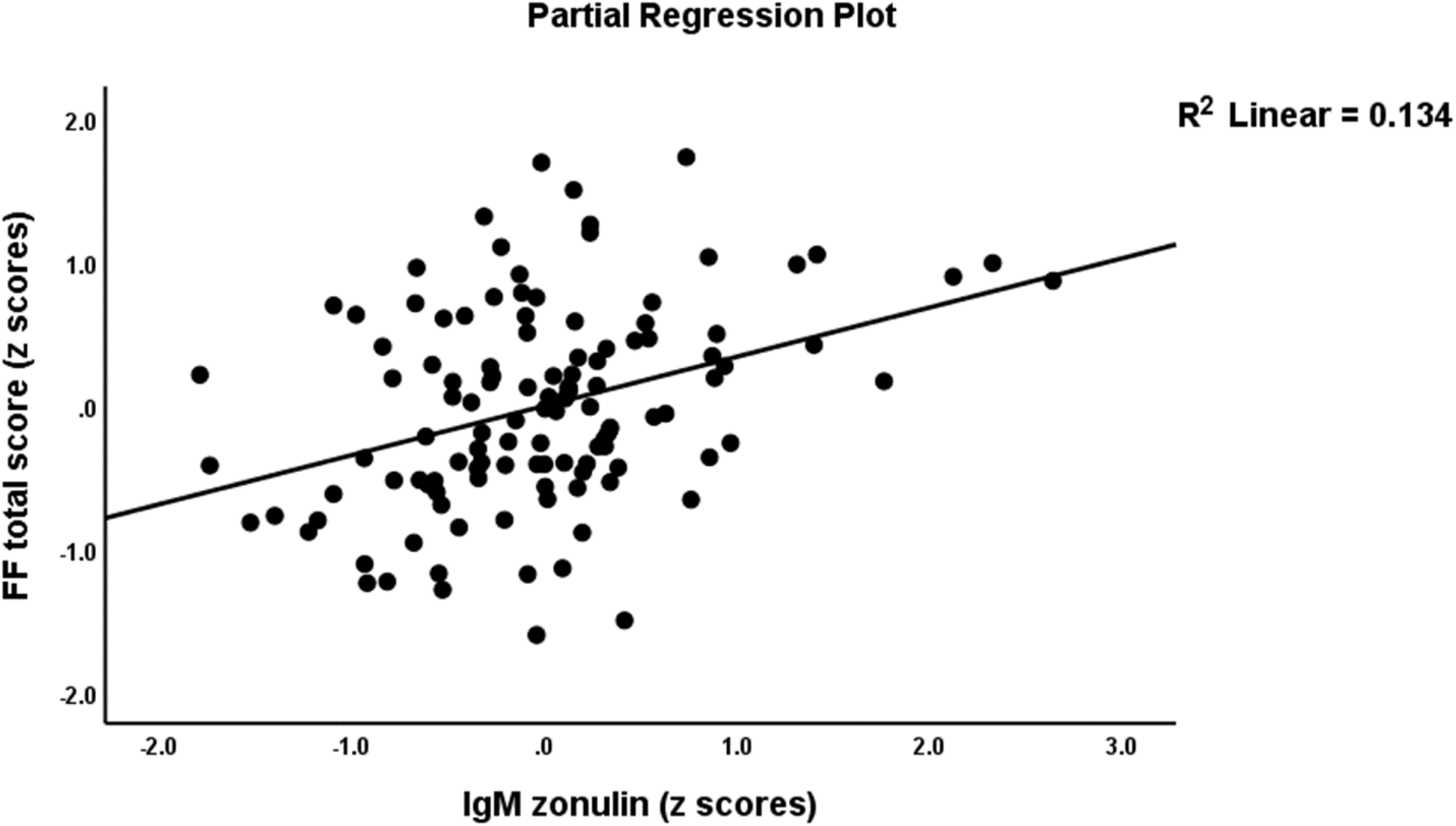
Partial regression of the total score on the Fibromyalgia and Chronic Fatigue rating scale on IgM levels to zonulin.

Table 3, regression #3 shows that 38.5% of the variance in FF physiosomatic score was explained by the regression on IgM zonulin, IgM PARA/TRANS, CCL11, IgA TRYCATs (all positively), and IgM protectome (inversely). Entering the G-CoDe in the same regression (regression #4) showed that 32.1% of the variance in the FF physiosomatic score was explained by the regression on G-CoDe, IgM zonulin, and IgA TRYCATs (all positively), and IgM protectome (inversely associated).

### Associations with QoL phenomenome

**Table 4** shows the outcome of multiple regression analyses with the WHO-QoL PC as dependent variable and both FF scores, the symptomatome, G-Code and biomarkers as explanatory variables. Table 4, regression #1 shows that 50.3% of the variance in the WHO-QoL PC was explained by the total FF score, symptomatome, and G-CoDe. **Figure 2** shows the partial regression of the QoL phenomenome on the total FF score. The second regression shows that 50.1% in the variance in the WHO-QoL score was explained by FF physiosomatic score, symptomatome, and G-CoDe.

**Table 4.**
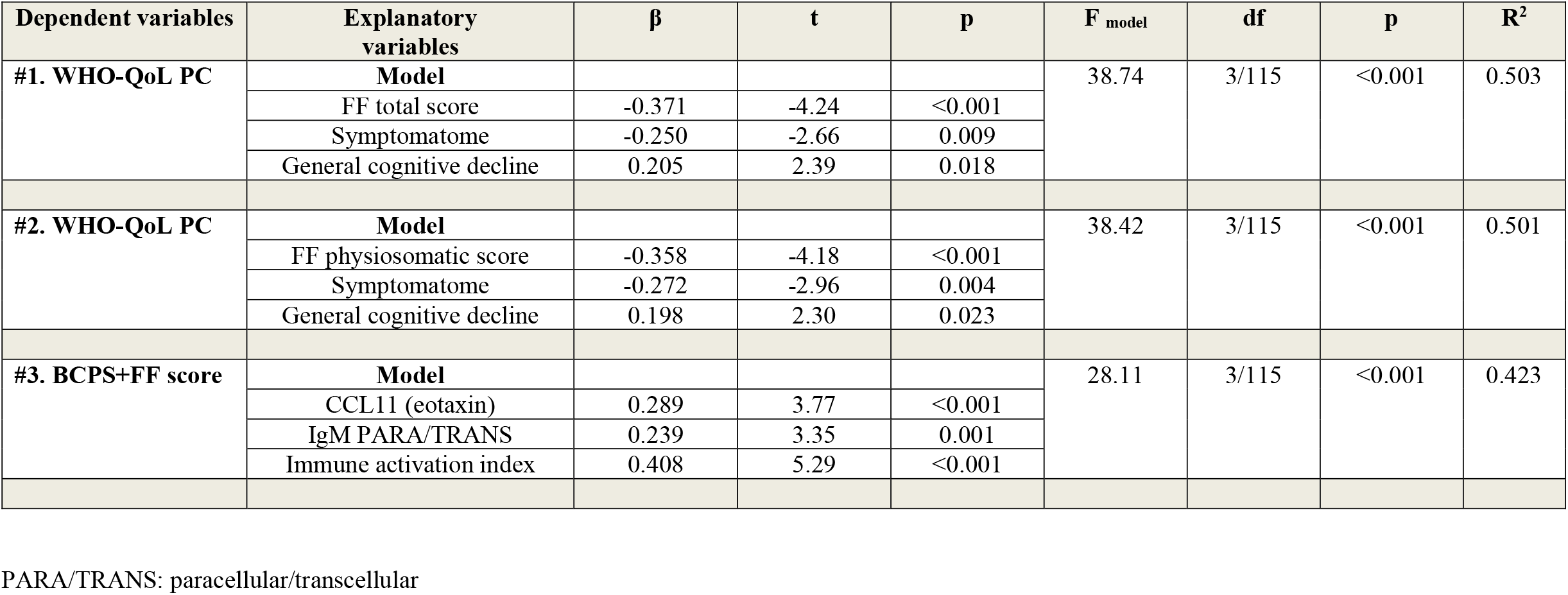
Results of multiple regression analyses with the WHO-QoL (quality of life) measurements or the first factor extracted from Fibromyalgia and Chronic Fatigue Rating (FF) scale score and the BCPS (behavioral-cognitive-physical-psychosocial)-worsening index (BCPS+FF) as dependent variables

**Figure 2.**
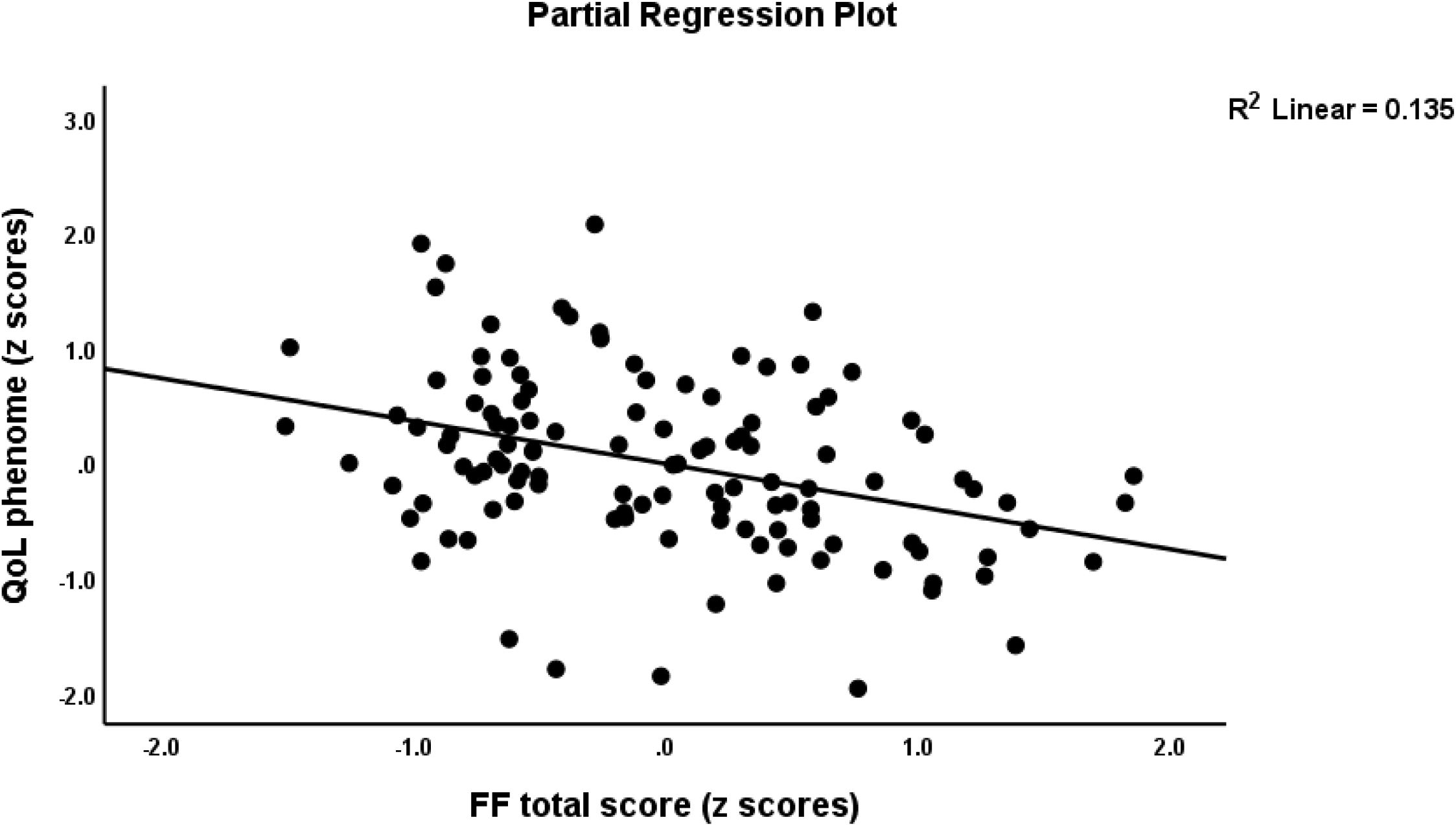
Partial regression of Quality of Life, phenomenome on the Fibromyalgia and Chronic Fatigue rating scale total score.

Given the strong associations between the FF scores and the other clinical scores (see Table 2), we have performed a PC analysis to determine whether the total FF score (and FF physiosomatic score) belong to the same PC as the symptomatome, G-CoDe, and WHO-Qol PC. We found that the first PC extracted from these 4 variables explained 69.8% of the variance and that all variables loaded highly on the first PC, namely total FF score: 0.840; symptomatome: 0.859; G-CoDe: −0.801; and WHO-Qol PC (−0.840). The KMO value (0.815) and Bartlett’s test of sphericity (X^2^=201.52, df=6, p<0.001) showed the factoriability of the data matrix was adequate. Using the FF physiosomatic score instead of the total FF score yielded comparable results, namely a loading of 0.830 on the first PC extracted from the 4 variables, which explained 69.3% of the variance and showed a KMO of 0.815. These results show that both FF scores belong to the same PC as the BCPS-worsening score. Consequently, we have computed the PC scores extracted from the total FF score and the three other clinical scores which reflects the BCBS + FF worsening index. Table 4, regression #3 shows that 42.3% of the variance in this BCBS+FF worsening score was explained by 3 biomarkers, namely CCL11, IgM PARA/TRANS, and immune activation index.

## Discussion

### Clinical aspects

The first major finding of this study is that schizophrenia is characterized by increased total FF scores and the severity of the physiosomatic symptoms of the FF and that the both FF scores are strongly associated with the BCPS-worsening index and its components, namely the symptomatome, G-CoDe, and QoL-phenomenome. Also, in other study groups, we found that FF symptoms as assessed with the FF scale, were part of the symptomatome of schizophrenia and belonged to a same latent factor as the PHEM and negative symptoms of schizophrenia (Almulla et al., 2020a; Mousa et al., 2020). This indicates that FF symptoms and physiosomatic symptoms are key manifestations of the symptomatome of schizophrenia. Moreover, the current study found that the total FF score and the severity of physiosomatic symptoms better predicted lowered quality of life that the symptomatome comprising PHEM and negative symptoms, PMR and FTD, and the G-CoDe as well.

Our finings that the total FF and physiosomatic FF scores were highly significantly associated with the BCPS-worsening index indicate that those FF symptoms are another feature of BCPS-worsening and should routinely be screened in schizophrenic patients to estimate the worsening of the illness. The results also suggest that FF symptoms and physiosomatic symptoms are driven, at least in part, by the same pathways that underpin aberrations in the connectome and neuroplasticity and which cause BCPS-worsening, namely “the prefronto-striato-thalamic, prefronto-parietal, prefronto-temporal, and dorsolateral prefrontal cortex, hippocampus and amygdala (Orellana et al., 2013; Orellana and Slachevsky, 2013; Maes et al., 2021b; Maes and Kanchanatawan, 2021).

### Biomarker correlates: focus on leaky gut

The second major finding of this study is that the total FF and physiosomatic FF scores are strongly associated with the IgM PARA/TRANS ratio and increased zonulin levels. Previously, we have described that the features of the symptomatome (all symptom domains) and cognitome (VFT, WLM, and True Recall) of schizophrenia were associated with the PARA/TRANS ratio indicating that increased gut permeability is associated with the phenome of schizophrenia (Maes et al., 2019a). Breakdown of the paracellular pathway and consequent increased bacterial translocation as indicated by increased IgA responses to Gram-negative bacteria are observed in deficit schizophrenia and especially in first-episode schizophrenia (Maes et al., 2021b). Interestingly, in the present study, IgM to zonulin was significantly associated with FF symptoms and physiosomatic symptoms, whereas previously no significant associations between zonulin and PHEM, negative symptoms, and memory deficits were detected in schizophrenia (Maes et al., 2019a). As a consequence, increased IgM to zonulin may be more specifically associated with physiosomatic symptoms which comprise gastro-intestinal symptoms (Maes et al., 2019a).

Zonulin increases gut permeability through effects on the tight junctions, which are composed of occludin and claudin (Vojdani and Vojdani, 2019). In physiological conditions, the tight junctions protect against translocation of Gram-negative bacteria, LPS, other microorganisms, and unwanted food allergens (Vojdani and Vojdani, 2021). Nevertheless, inflammation, viral infections, psychosocial stress, gut dysbiosis, and nutritional factors may induce loosening of the tight junctions, thereby causing translocation of bacterial LPS and other antigens into the blood stream via a leaky paracellular route (Vojdani and Vojdani, 2021; Rudzki and Maes, 2020; 2021). It is important to note that, in schizophrenia, not only the tight junctions, but also the adherens junctions and the vascular barrier may be dysfunctional (Maes et al., 2019b). Interestingly, zonulin or pre-haptoglobin-(Hp)-2 is associated with the Hp-2 gene (Maes et al., 2001) and genetic polymorphism of the Hp gene on chromosome 16 determines the Hp 1-1, Hp 2-1 and Hp 2-2 phenotypes whereby the latter is characterized by increased inflammatory responses with elevated levels of interleukin (IL)-6 and tumor necrosis factor (TNF)-α and oxidative stress (Maes et al., 1994; 2001). Moreover, it is important to note that the plasma levels of Hp, which is an acute phase protein (APP), are significantly increased in schizophrenia in conjunction with other acute phase proteins and complement factors, including C3C (Maes et al., 1997). As such, part of the immune-inflammatory and oxidative stress responses in schizophrenia (Maes et al., 1997; 2021b) may be explained by an increased Hp-2 gene frequency, increased zonulin levels and accompanying leaky gut (Maes et al., 2001; 2019a).

It is interesting to note that the severity of FF symptoms in ME/CFS is significantly associated with indicants of leaky gut including increased IgA and IgM responses to the same Gram-negative bacteria as determined in the present study (Maes et al., 2007). Moreover, treatment with specific antioxidants, which are known to improve leaky gut (zinc, glutamine, and N-acetyl-cysteine) improves these indicants of leaky gut in association with a significant improvement in the severity of FF symptoms (Maes and Leunis, 2008). Part of the schizophrenia patients, especially those with first episode schizophrenia, show leaky gut and increased bacterial translocation an, therefore, those patients may benefit from treatments for leaky gut (Maes et al., 2021b).

### Biomarker correlates: focus on immune activation and neurotoxicity

The third major finding of the present study is that a considerable part of the variance in the total FF score and the severity of physiosomatic FF symptoms is explained not only by the PARA/TRANS ratio and IgM to zonulin, but also by immune activation as indicated by increased levels of IL-6, TNF-α, MIP-1, and IL-10, and CCL11, and by lowered IgM responses to OSEs. These cytokines/chemokines are all increased during an immune-inflammatory response with IL-6 and TNF-α displaying inflammatory, CCL11 and MIP-1 chemotaxic, and IL-10 immune-regulatory effects (Maes et al., 2021b). These results extend those of previous reports which showed that a larger part of the variance in the FF score in schizophrenia is accompanied by increased levels of CCL11 and high mobility group box 1 protein (HMGB1) (Mousa et al., 2020). HMGB1 is a DAMP (damage-associated molecular pattern) that may be released by necrotic and injured cells and stimulates the production and release of IL-6 and TNF-α, and T helper-(Th)-1 cytokines including interferon-γ (Yang et al., 2016; Lotze et al., 2005; Festoff et al., 2016). The results of the current study also extend those of Almulla et al. (2020a) who reported that FF scores in schizophrenia are associated with increased TNF-α and CCL11 and with indicants of IL-1 signaling, namely increased IL-1β and sIL-1RA levels. Moreover, these immune biomarkers explained 59.4% of the variance in a factor extracted from the FF and schizophrenia symptoms (Almulla et al., 2020a). In both studies, HMGB1, IL-6, CLL11, TNF-α, CCL11, IL-1β, and/or sIL-1RA significantly predicted cognitive deficits including in executive functions, semantic and episodic memory, and attention (Almulla et al., 2020a; Al-Dujaili et al., 2021). Importantly, a large part (45.1%) of the variance in the first factor extracted from the symptomatome, cognitome and the total FF score was explained by HMGB1 and CCL11 levels coupled with changes in the endogenous opioid system (Mousa et al., 2021). Furthermore, the severity of FF symptoms was also associated with lowered IgM responses to OSEs and the latter also predicted the cognitome and symptomatome of schizophrenia (Maes et al., 2020b).

It is important to note that the severity of FF symptoms in ME/CFS is significantly associated with signs of immune activation including increased levels of pro-inflammatory cytokines, elastase, and lysozyme, oxidative damage, hypernitrosylation, mitrochondrial disorders, and changes in the gut-brain axis (Maes and Twisk, 2010; Maes et al., 2012; Morris and Maes, 2014a; Hornig et al., 2017; Bjorklund et al., 2020; Strawbridge et al., 2019; Nagy-Szakal et al., 2017; Anderson and Maes, 2020; Lupo et al., 2021). There is also some evidence that fibromyalgia is accompanied by immune activation as reviewed in a recent meta-analysis, which showed increased IL-6, IL-17, and IL-4 levels, and increased M1 and T regulatory profiles, although the effect sizes were very modest (Andrés-Rodríguez et al., 2020).

Interestingly, the combined impact of leaky gut, immune activation and natural IgM biomarkers was more important than that of neurotoxicity indices, although the MITOTOX index comprising the neurotoxic cytokines/chemokines and TRYCATs coupled with LPS and the oxidative stress damage was strongly associated with the physiosomatic symptoms. Previously, we have discussed that the symptomatome, G-CoDe, Qol-phenome, and the BCPS-worsening index are significantly associated with indices of increased neurotoxicity as indicated by increased levels of neurotoxic cytokines including IL-6, TNF-α, CCL11, MIP-1, increased levels of neurotoxic TRYCATs, including PA, XA, and 3HK, and signs of oxidative/chlorinative stress as indicated by increased LOOH, MDA, and AOPP levels (Maes et al., 2020a; 2021b).

### Mechanistic explanations

This study showed that FF symptoms belong to the same common core shared by G-CoDe, and the symptomatome and, therefore, we may suggest that at least part of the variability in FF symptoms is determined by the same pathways which underpin cognitive deficits and the symptomatic phenome. The mechanistic explanation is that the symptomatome and cognitome of schizophrenia are probably caused by pathways which cause alterations in gray and white matter which lead to aberrations in neuroplasticity and connectivity especially in the salience network (anterior insula and anterior cingulate cortex), amygdala, the prefronto-striato-thalamic, prefronto-parietal, prefronto-temporal, and dorsolateral prefrontal cortex (Orellana et al., 2013; Orellana and Slachevsky, 2013; Maes et al., 2020a; 2021b; Stoyanov and Maes, 2021; Stoyanov et al., 2021). We have described previously how these same peripheral pathways including leaky gut, immune activation, TRYCAT pathway activation, and nitro-oxidative stress may cause central changes leading to physiosomatic symptoms such as central fatigue and a flu-like malaise, and affective, cognitive and autonomic symptoms (Maes and Twisk, 2010; Morris and Maes, 2013; 2014b; Morris et al., 2016; 2019). Nevertheless, effects of peripheral immune-inflammatory pathways on peripheral tissues including muscle, gut and the cardio-vascular system may additionally contribute to muscle pain and spams, and gastro-intestinal and cardio-vascular symptoms (Morris and Maes, 2014b; Morris et al., 2016; Jonsjö et al., 2019). Furthermore, natural IgM responses to OSEs are part of the innate immune system and are a first-line defense against increased pathogen invasion, including Gram-negative bacteria, inflammation and oxidative stress (Maes et al., 2021b). As such, lowered natural IgM responses to OSEs may predispose towards increased immune responses and nitro-oxidative stress (Maes et al., 2020b) and, thus, to the onset of FF symptoms in schizophrenia.

### Limitations

Our results should be interpreted with regard to its limitations. Firstly, this is a case-control study, and this design does not allow to draw solid conclusions on causality. Secondly, it would have been more interesting if we had measured M1, Th-1, Th-2, and T regulatory profiles as well as the connectome using functional MRI.

## Conclusions

The combined effects of leaky gut, immune and TRYCAT pathway activation and lowered protection due to lowered natural IgM levels drive in part the physiosomatic symptoms in schizophrenia. Physiosomatic symptoms are part of the phenome of schizophrenia and the BCPS-worsening index, and they strongly predict a lowered quality of life.

## Data Availability

The dataset generated during and/or analyzed during the current study will be available from the corresponding author upon reasonable request and once the dataset has been fully exploited by the authors.

## Compliance with ethical standards

### Research involving human participants

Approval for the study was obtained from the Institutional Review Board of Chulalongkorn University, Bangkok, Thailand.

### Informed consent

All controls and patients and the guardians of patients gave written informed consent before participation in our study.

### Funding

This research has been supported by the Asahi Glass Foundation, Chulalongkorn University Centenary Academic Development Project.

### Author’s contributions

All the contributing authors have participated in the manuscript. BK and MM designed the study. BK recruited patients and completed diagnostic interviews and rating scales measurements. MM carried out the statistical analyses. AV, SS and DB performed the analyses. All authors contributed to the interpretation of the data and writing of the manuscript.

## References

Al-Dujaili, A.H., Mousa, R.F., Al-Hakeim, H.K., Maes, M., 2021. High Mobility Group Protein 1 and Dickkopf-Related Protein 1 in Schizophrenia and Treatment-Resistant Schizophrenia: Associations With Interleukin-6, Symptom Domains, and Neurocognitive Impairments. Schizophr. Bull. 47(2), 530-541. doi:10.1093/schbul/sbaa136. PMID: 32971537; PMCID: PMC7965081.

Al-Hakeim, H.K., Almulla, A.F., Al-Dujaili, A.H., Maes, M., 2020. Construction of a Neuro-Immune-Cognitive Pathway-Phenotype Underpinning the Phenome of Deficit Schizophrenia. Curr. Top. Med. Chem. 20(9), 747-758. doi:10.2174/1568026620666200128143948. PMID: 31994463.

Almulla, A.F., Al-Hakeim, H.K., Abed, M.S., Carvalho, A.F., Maes, M., 2020a. Chronic fatigue and fibromyalgia symptoms are key components of deficit schizophrenia and are strongly associated with activated immune-inflammatory pathways. Schizophr Res. 222, 342-353. doi:10.1016/j.schres.2020.05.003. Epub 2020 May 26. PMID: 32467068.

Almulla, A.F., Al-Hakeim, H.K., Maes, M., 2020b. Schizophrenia phenomenology revisited: positive and negative symptoms are strongly related reflective manifestations of an underlying single trait indicating overall severity of schizophrenia. CNS Spectr. 20, 1-10. doi:10.1017/S1092852920001182. Epub ahead of print. PMID: 32431263.

Anderson, G., Maes, M., 2020. Mitochondria and immunity in chronic fatigue syndrome. Prog. Neuropsychopharmacol. Biol. Psychiatry. 103, 109976. doi:10.1016/j.pnpbp.2020.109976. Epub 2020 May 26. PMID: 32470498.

Andreasen, N.C., 1989. The Scale for the Assessment of Negative Symptoms (SANS): conceptual and theoretical foundations. Brit. J. Psychiatry Suppl. 7, 49–58.

Andrés-Rodríguez, L., Borràs, X., Feliu-Soler, A., Pérez-Aranda, A., Angarita-Osorio, N., Moreno-Peral, P., Montero-Marin, J., García-Campayo, J., Carvalho, A.F., Maes, M., Luciano, J.V., 2020. Peripheral immune aberrations in fibromyalgia: A systematic review, meta-analysis and meta-regression. Brain Behav. Immun. 87, 881-889. doi:10.1016/j.bbi.2019.12.020. Epub 2019 Dec 27. PMID: 31887417.

Bjørklund, G., Dadar, M., Pivina, L., Doşa, M.D., Semenova, Y., Maes, M., 2020. Environmental, Neuro-immune, and Neuro-oxidative Stress Interactions in Chronic Fatigue Syndrome. Mol. Neurobiol. 57(11), 4598-4607. doi:10.1007/s12035-020-01939-w. Epub 2020 Aug 6. PMID: 32761353.

CANTAB, 2018. The most validated cognitive research software. http://www.cambridgecognition.com/cantab/ October 1.

CERAD, 1986. CERAD – An Overview: The Consortium to Establish a Registry for Alzheimer’s Disease. http://cerad.mc.duke.edu/

Festoff, B.W., Sajja, R.K., van Dreden, P., Cucullo, L., 2016. HMGB1 and thrombin mediate the blood-brain barrier dysfunction acting as biomarkers of neuroinflammation and progression to neurodegeneration in Alzheimer’s disease. J. Neuroinflammation 13, 194.

Hamilton, M., 1960. A rating scale for depression. J. Neurol. Neurosurg. Psychiatry. 23, 56–62.

Hornig, M., Montoya, J.G., Klimas, N.G., Levine, S., Felsenstein, D., Bateman, L., Peterson, D.L., Gottschalk, C.G., Schultz, A.F., Che, X., Eddy, M.L., Komaroff, A.L., Lipkin, W.I., 2015. Distinct plasma immune signatures in ME/CFS are present early in the course of illness. Sci. Adv. 1(1), e1400121. doi:10.1126/sciadv.1400121. PMID: 26079000; PMCID: PMC4465185.

Jonsjö, M.A., Olsson, G.L., Wicksell, R.K., Alving, K., Holmström, L., Andreasson, A., 2020. The role of low-grade inflammation in ME/CFS (Myalgic Encephalomyelitis/Chronic Fatigue Syndrome) - associations with symptoms. Psychoneuroendocrinology. 113, 104578. doi:10.1016/j.psyneuen.2019.104578. Epub 2019 Dec 26. PMID: 31901625.

Kanchanatawan, B., Sirivichayakul, S., Thika, S., Ruxrungtham, K., Carvalho, A.F., Geffard, M., Anderson, G., Noto, C., Ivanova, R., Maes, M., 2017. Physio-somatic symptoms in schizophrenia: association with depression, anxiety, neurocognitive deficits and the tryptophan catabolite pathway. Metab. Brain Dis. 32(4), 1003-1016. doi:10.1007/s11011-017-9982-7. Epub 2017 Mar 3. PMID: 28258445.

Kanchanatawan, B., Sriswasdi, S., Maes, M., 2019. Supervised machine learning to decipher the complex associations between neuro-immune biomarkers and quality of life in schizophrenia. Metab. Brain Dis.34(1), 267-282. doi:10.1007/s11011-018-0339-7. Epub 2018 Nov 22. PMID: 30467771.

Kay, S.R., Fiszbein, A., Lindenmayer, J.P., et al., 1986. Positive and negative syndromes in schizophrenia as a function of chronicity. Acta Psychiatr. Scand. 74(5), 507-518. doi:10.1111/j.1600-0447.1986.tb06276.x. PMID: 3492863.

Kittirathanapaiboon, P., Khamwongpin, M., 2005. The Validity of the Mini International Neuropsychiatric Interview (M.I.N.I.) Thai Version: Suanprung Hospital, Department of Mental Health.

Lotze, M.T., Tracey, K.J., 2005. High-mobility group box 1 protein (HMGB1): Nuclear weapon in the immune arsenal. Nat. Rev. Immunol. 5, 331–342.

Lupo, G.F.D., Rocchetti, G., Lucini, L., Lorusso, L., Manara, E., Bertelli, M., Puglisi, E., Capelli, E., 2021. Potential role of microbiome in Chronic Fatigue Syndrome/Myalgic Encephalomyelits (CFS/ME). Sci Rep. 11(1), 7043. doi:10.1038/s41598-021-86425-6. PMID: 33782445; PMCID: PMC8007739.

Maes, M., Leunis, J.C., 2008. Normalization of leaky gut in chronic fatigue syndrome (CFS) is accompanied by a clinical improvement: effects of age, duration of illness and the translocation of LPS from gram-negative bacteria. Neuro Endocrinol. Lett. 29(6), 902–910. PMID: 19112401.

Maes, M., Twisk, F.N., 2010. Chronic fatigue syndrome: Harvey and Wessely’s (bio)psychosocial model versus a bio(psychosocial) model based on inflammatory and oxidative and nitrosative stress pathways. BMC Med. 8, 35. doi:10.1186/1741-7015-8-35. PMID: 20550693; PMCID: PMC2901228.

Maes, M., Kanchanatawan, B., 2021. In (deficit) schizophrenia, a general cognitive decline partly mediates the effects of neuro-immune and neuro-oxidative toxicity on the symptomatome and quality of life. CNS Spectr. 12, 1-10. doi:10.1017/S1092852921000419. Epub ahead of print. PMID: 33843548.

Maes, M., Delanghe, J., Scharpé, S., Meltzer, H.Y., Cosyns, P., Suy, E., Bosmans, E., 1994. Haptoglobin phenotypes and gene frequencies in unipolar major depression. Am. J. Psychiatry. 151(1), 112-116. doi:10.1176/ajp.151.1.112. PMID: 8267107.

Maes, M., Delange, J., Ranjan, R., Meltzer, H.Y., Desnyder, R., Cooremans, W., Scharpé, S., 1997; Acute phase proteins in schizophrenia, mania and major depression: modulation by psychotropic drugs. Psychiatry Res. 66(1), 1-11. doi:10.1016/s0165-1781(96)02915-0. PMID: 9061799.

Maes, M., Delanghe, J., Bocchio Chiavetto, L., Bignotti, S., Tura, G.B., Pioli, R., Zanardini, R., Altamura, C.A., 2001. Haptoglobin polymorphism and schizophrenia: genetic variation on chromosome 16. Psychiatry Res. 104(1), 1-9. doi:10.1016/s0165-1781(01)00298-0. PMID: 11600184.

Maes, M., Mihaylova, I., Leunis, J.C., 2007. Increased serum IgA and IgM against LPS of enterobacteria in chronic fatigue syndrome (CFS): indication for the involvement of gram-negative enterobacteria in the etiology of CFS and for the presence of an increased gut-intestinal permeability. J. Affect. Disord. 99(1-3), 237-240. doi:10.1016/j.jad.2006.08.021. Epub 2006 Sep 27. PMID: 17007934.

Maes, M., Twisk, F.N., Kubera, M., Ringel, K., 2012. Evidence for inflammation and activation of cell-mediated immunity in Myalgic Encephalomyelitis/Chronic Fatigue Syndrome (ME/CFS): increased interleukin-1, tumor necrosis factor-α, PMN-elastase, lysozyme and neopterin. J. Affect. Disord. 136(3), 933-939. doi:10.1016/j.jad.2011.09.004. Epub 2011 Oct 4. PMID: 21975140.

Maes, M., Sirivichayakul, S., Kanchanatawan, B., Vodjani, A., 2019a. Upregulation of the Intestinal Paracellular Pathway with Breakdown of Tight and Adherens Junctions in Deficit Schizophrenia. Mol. Neurobiol. 56(10), 7056-7073. doi:10.1007/s12035-019-1578-2. Epub 2019 Apr 10. PMID: 30972627.

Maes, M., Sirivichayakul, S., Kanchanatawan, B., Vodjani, A., 2019b. Breakdown of the Paracellular Tight and Adherens Junctions in the Gut and Blood Brain Barrier and Damage to the Vascular Barrier in Patients with Deficit Schizophrenia. Neurotox. Res. 36(2), 306-322. doi:10.1007/s12640-019-00054-6. Epub 2019 May 10. PMID: 31077000.

Maes, M., Sirivichayakul, S., Matsumoto, A.K., Michelin, A.P., de Oliveira Semeão, L., de Lima Pedrão, J.V., Moreira, E.G., Barbosa, D.S., Carvalho, A.F., Solmi, M., Kanchanatawan, B., 2000a. Lowered Antioxidant Defenses and Increased Oxidative Toxicity Are Hallmarks of Deficit Schizophrenia: a Nomothetic Network Psychiatry Approach. Mol. Neurobiol. 57(11), 4578-4597. doi:10.1007/s12035-020-02047-5. Epub 2020 Aug 5. PMID: 32754898.

Maes, M., Vojdani, A., Galecki, P., Kanchanatawan, B,.2020b. How to Construct a Bottom-Up Nomothetic Network Model and Disclose Novel Nosological Classes by Integrating Risk Resilience and Adverse Outcome Pathways with the Phenome of Schizophrenia. Brain Sci. 10(9), 645. doi:10.3390/brainsci10090645. PMID: 32957709; PMCID: PMC7565440.

Maes, M., Moraes, J.B., Bonifacio, K.L., Barbosa, D.S., Vargas, H.O., Michelin, A.P., Nunes, S.O.V., 2021a. Towards a new model and classification of mood disorders based on risk resilience, neuro-affective toxicity, staging, and phenome features using the nomothetic network psychiatry approach. Metab. Brain Dis. 36(3), 509-521. doi:10.1007/s11011-020-00656-6. Epub 2021a Jan 7. PMID: 33411213.

Maes, M., Vojdani, A., Sirivichayakul, S., Barbosa, D.S., Kanchanatawan, B., 2021b. Inflammatory and Oxidative Pathways Are New Drug Targets in Multiple Episode Schizophrenia and Leaky Gut, Klebsiella pneumoniae, and C1q Immune Complexes Are Additional Drug Targets in First Episode Schizophrenia. Mol. Neurobiol. Mar 6. doi:10.1007/s12035-021-02343-8. Epub ahead of print. PMID: 33675500.

Morris, G., Maes, M., 2013. Myalgic encephalomyelitis/chronic fatigue syndrome and encephalomyelitis disseminata/multiple sclerosis show remarkable levels of similarity in phenomenology and neuroimmune characteristics. BMC Med. 11, 205. doi:10.1186/1741-7015-11-205. PMID: 24229326; PMCID: PMC3847236.

Morris, G., Maes, M., 2014a. Oxidative and Nitrosative Stress and Immune-Inflammatory Pathways in Patients with Myalgic Encephalomyelitis (ME)/Chronic Fatigue Syndrome (CFS). Curr. Neuropharmacol. 12(2), 168-185. doi:10.2174/1570159X11666131120224653. PMID: 24669210; PMCID: PMC3964747.

Morris, G., Maes, M., 2014b. Mitochondrial dysfunctions in myalgic encephalomyelitis/chronic fatigue syndrome explained by activated immuno-inflammatory, oxidative and nitrosative stress pathways. Metab. Brain Dis. 29(1), 19-36. doi:10.1007/s11011-013-9435-x. Epub 2013 Sep 10. PMID: 24557875.

Morris, G., Berk, M., Galecki, P., Walder, K., Maes, M., 2016. The Neuro-Immune Pathophysiology of Central and Peripheral Fatigue in Systemic Immune-Inflammatory and Neuro-Immune Diseases. Mol. Neurobiol. 53(2), 1195-1219. doi:10.1007/s12035-015-9090-9. Epub 2015 Jan 20. PMID: 25598355.

Morris, G., Puri, B.K., Walker, A.J., Maes, M., Carvalho, A.F., Walder, K., Mazza, C., Berk, M., 2019. Myalgic encephalomyelitis/chronic fatigue syndrome: From pathophysiological insights to novel therapeutic opportunities. Pharmacol. Res. 148, 104450. doi:10.1016/j.phrs.2019.104450. Epub 2019 Sep 8. PMID: 31509764.

Mousa, R.F., Al-Hakeim, H.K., Alhaideri, A., Maes, M., 2021. Chronic fatigue syndrome and fibromyalgia-like symptoms are an integral component of the phenome of schizophrenia: neuro-immune and opioid system correlates. Metab. Brain Dis. 36(1), 169-183. doi:10.1007/s11011-020-00619-x. Epub 2020 Sep 23. PMID: 32965599.

Nagy-Szakal, D., Williams, B.L., Mishra, N., Che, X., Lee, B., Bateman, L., Klimas, N.G., Komaroff, A.L., Levine, S., Montoya, J.G., Peterson, D.L., Ramanan, D., Jain, K., Eddy, M.L., Hornig, M., Lipkin, W.I., 2017. Fecal metagenomic profiles in subgroups of patients with myalgic encephalomyelitis/chronic fatigue syndrome. Microbiome. 5(1), 44. doi:10.1186/s40168-017-0261-y. PMID: 28441964; PMCID: PMC5405467.

Orellana, G., Slachevsky, A., 2013. Executive functioning in schizophrenia. Front Psychiatry. 4, 1–15.

Orellana, G., Alvarado, L., Muñoz-Neira, C., et al., 2013. A. Psychosis-related matricide associated with a lesion of the ventromedial prefrontal cortex. J. Am. Acad. Psychiatry Law 41(3), 401–406.

Overall, J.E., Gorham, D.R., 1962. The brief psychiatric rating scale. Psychological Reports. 10, 799–812.

Strawbridge, R., Sartor, M.L., Scott, F., Cleare, A.J., 2019. Inflammatory proteins are altered in chronic fatigue syndrome-A systematic review and meta-analysis. Neurosci. Biobehav. Rev. 107, 69-83. doi:10.1016/j.neubiorev.2019.08.011. Epub 2019 Aug 26. PMID: 31465778.

Roomruangwong, C., Kanchanatawan, B., Carvalho, A.F., Sirivichayakul, S., Duleu, S., Geffard, M., Maes, M., 2018. Body image dissatisfaction in pregnant and non-pregnant females is strongly predicted by immune activation and mucosa-derived activation of the tryptophan catabolite (TRYCAT) pathway. World J Biol Psychiatry. 19(3), 200-209. doi:10.1080/15622975.2016.1213881. Epub 2016 Aug 30. PMID: 27427239.

Rudzki, L., Maes, M., 2020. The Microbiota-Gut-Immune-Glia (MGIG) Axis in Major Depression. Mol. Neurobiol. 57(10), 4269-4295. doi:10.1007/s12035-020-01961-y. Epub 2020 Jul 22. PMID: 32700250.

Rudzki, L., Maes, M., 2021. From “Leaky Gut” to Impaired Glia-Neuron Communication in Depression. Adv. Exp. Med. Biol. 1305, 129-155. doi:10.1007/978-981-33-6044-0_9. PMID: 33834399.

Stoyanov, D., Maes, M.H., 2021a. How to construct neuroscience-informed psychiatric classification? Towards nomothetic networks psychiatry. World J. Psychiatry. 11(1), 1-12. doi:10.5498/wjp.v11.i1.1. PMID: 33511042; PMCID: PMC7805251.

Stoyanov, D., Aryutova, K., Kandilarova, S., Paunova, R., Arabadzhiev, Z., Todeva-Radneva, A., Kostianev, S., Borgwardt, S., 2021b. Diagnostic Task Specific Activations in Functional MRI and Aberrant Connectivity of Insula with Middle Frontal Gyrus Can Inform the Differential Diagnosis of Psychosis. Diagnostics (Basel). 11(1), 95. doi:10.3390/diagnostics11010095. PMID: 33435624; PMCID: PMC7827259.

Vojdani, A., Vojdani, E., 2019. Food-associated autoimmunities: when food breaks your immune system (autoimmunity). A&g Wilshire, LLC, pp 454. ISBN-13: 978-0578499772

WHO, 1993. Study protocol for the World Health Organization project to develop a Quality of Life assessment instrument (WHOQOL). Qual, Life Res. 2(2), 153–159.

Yang, H., Wang, H., Levine, Y.A, et al., 2016. Identification of CD163 as an anti-inflammatory receptor for HMGB1-haptoglobin complexes. JCI Insight 1, e85375.

Zachrisson, O., Regland, B., Jahreskog, M., Kron, M., Gottfries, C.G., 2002. A rating scale for fibromyalgia and chronic fatigue syndrome (the FibroFatigue scale) J. Psychosom. Res. 52, 501–509.

